# Exploring the Relationship Between Apathy, Dopaminergic Signal, and Head Injury in Neurodevelopmental Disorders

**DOI:** 10.64898/2026.05.14.26353215

**Authors:** Rubina Malik, Sarah Al-Saoud, Kassandra Rogers, Emma G. Duerden

## Abstract

Apathy is characterized by reduced motivation for goal-directed behaviour and may emerge following brain injury. Currently, little is known about apathy in children and adolescents with neurodevelopmental disorders (NDDs) exposed to repetitive head impacts. This exploratory study investigated associations between apathy, repetitive head-banging behaviour, and substantia nigra neuromelanin-sensitive MRI (NM-MRI) signal in youth with NDDs. Forty-seven participants (14 typically developing; 33 ADHD/ASD) completed Behaviour Assessment System for Children (BASC-3) measures, from which apathy-related items were harmonized across developmental forms and subjected to principal component analysis. A one-component solution explained 47.3% of variance and was used to derive apathy scores. Although head-banging severity and NM-MRI signal were not independently associated with apathy, a significant interaction emerged, whereby greater head-banging severity strengthened the relationship between apathy and substantia nigra NM-MRI signal. These preliminary findings suggest repetitive self-injurious head impacts may influence dopaminergic systems linked to motivational dysfunction in youth with NDDs.

## 1. INTRODUCTION

Apathy is a neuropsychiatric syndrome characterized by reduced motivation for goal-directed behaviour and cognition. ^1–3^ It can manifest across multiple domains, including social engagement, emotional responsiveness, and behavioural activation.^1^ Apathy is a common outcome of traumatic brain injury (TBI) in adults, with an estimated prevalence of approximately 47%,^4^ and is associated with poorer functional outcomes, including reduced daily functioning, delayed recovery, cognitive impairment^5^, and increased caregiver burden.^6^ Moreover, apathy following repetitive TBIs has been linked to chronic traumatic encephalopathy (CTE), a neurodegenerative condition that can lead to dementia in severe cases.^7^ Despite its clinical significance, there is currently no instrument specifically developed or validated to assess apathy in TBI populations.^4^

Children and adolescents with neurodevelopmental disorders (NDDs), including attention-deficit/hyperactivity disorder (ADHD) and autism spectrum disorder (ASD), are at substantially increased risk of sustaining TBIs compared to their neurotypically developing peers.^8–10^ Contributing factors include impulsivity, exposure to violence, and self-injurious behaviours (SIB), particularly head-banging. Head-banging is a stereotyped motor behaviour involving repetitive impact of the head against surfaces, often emerging in early childhood and, in some cases, persisting into adolescence.^11^ While repetitive head impacts are a known risk factor for brain injury, the relationship between head-banging and TBI in youth with NDDs remains poorly understood. Limited evidence, including case reports of individuals with long-standing head-banging behaviour, suggests the potential for neuropathological changes consistent with repetitive brain injury^12–14^; however, systematic research in pediatric populations is lacking.

Despite the elevated risk of TBI in youth with NDDs, apathy following TBI is not well characterized in this group. More broadly, the developmental trajectory of apathy remains unclear, in part due to the lack of standardized, age-appropriate measurement tools.^15^ Emerging work suggests that apathy is associated with structural brain changes following pediatric TBI, including reduced grey and white matter volume.^16^ Other studies have used behavioural paradigms, such as reward-based decision-making tasks, to index motivational deficits. In adolescents (ages 12–18 years old) with chronic TBI (> 1 year post-injury) a motivation Go/No-Go task that rewarded correct responses and punished incorrect responses was employed; compared to controls, the adolescents with TBI demonstrated lower fronto-striatal connectivity that correlated with poorer Motivation task performance.^17^ Similarly, children ages 7–15 years with TBI have been shown to perform more poorly on a decision-making task, compared to controls.^18^ Together, these findings suggest that disruptions in motivational systems may persist following pediatric TBI and contribute to functional impairment.

An emerging technique in pediatric neuroimaging is neuromelanin-sensitive magnetic resonance imaging (NM-MRI), which provides an *in vivo* measurement of catecholaminergic systems integrity by quantifying neuromelanin accumulation within the substantia nigra (SN) dopaminergic neurons.^19^ Dopaminergic systems are known to underlie motivation and goal-directed behaviour,^20^ making NM-MRI particularly well suited for investigating the neurobiological basis of apathy. In pediatric populations, NM-MRI offers a feasible and sensitive method for detecting early alterations in these systems, enabling the study of brain–behavior relationships across neurodevelopmental and clinical groups.

In the absence of a gold-standard measure of apathy for children and adolescents, dimensional approaches that leverage existing behavioural instruments may provide a useful alternative. In the present exploratory study, we operationalized apathy using a set of theoretically derived items from the Behavior Assessment System for Children (BASC-3), harmonized across developmental versions to capture a common motivational construct. We then examined whether this measure of apathy was associated with the severity of head-banging behaviour, given its potential role as a source of repetitive head injury in youth with NDDs.

Building on evidence implicating dopaminergic systems in motivation, we further investigated whether apathy was related to NM-MRI signal intensity within the substantia nigra (SN-NM), and whether head-banging severity moderated this relationship. Specifically, we tested (1) whether greater head-banging severity was associated with higher levels of apathy, (2) whether greater SN-NM signal change was associated with greater apathy, and (3) whether the relationship between SN-NM signal and apathy varied as a function of head-banging severity. Given the modest sample size, these analyses were exploratory and intended to generate preliminary evidence for a mechanistic link between repetitive head injury, dopaminergic system integrity, and apathy across development.

## 2. METHODS

### 2.1 Participants

Children and adolescents between the ages of 4 and 21 years were included in this study. Participants were recruited between January 2020–December 2025 from community advertisements in London, Ontario, Canada. The sample included typically developing (TD) youth and youth with a clinical diagnosis of ADHD or ASD, grouped as participants with neurodevelopmental disorders (NDD). This research was approved by the Health Sciences Research Ethics Board at Western University. Participants provided written informed consent or assent, or parental consent for participants <18 years.

### 2.2 Questionnaires

Head-banging severity was assessed using caregiver-report, age-appropriate measures of restrictive and repetitive behaviours (Repetitive Behaviors Scale–Revised; Repetitive Behavior Scale–Early Childhood), which include a self-injurious behaviour (SIB) domain with similar item content and scoring structure. Within the SIB data, three items pertaining to head-banging were extracted and combined as a proxy for repetitive head injury severity, SIB-head: 1) Hits or slaps head, face, or other body area; 2) Hits or bangs head or other body part on table, floor or other surface; 3) Hits or bangs head or other body area with objects. Each item is rated on a 4-point severity scale (0–3) to identify patterns of stereotyped and self-injurious behaviour.

The Behavior Assessment System for Children, Third Edition (BASC-3; child and adolescent forms) was used to assess symptoms related to apathy. Items reflecting motivational, emotional, and executive deficits were extracted (BASC-3-apathy; see table S1 for extracted items), and used in a principal component analysis (PCA) to identify apathy constructs present in the sample.

The full-scale IQ (FSIQ), used to assess general intelligence, is derived from standardized tests, such as the Wechsler scales for children. It represents overall cognitive ability across domains, including verbal comprehension, reasoning, working memory, and processing speed. The FSIQ was used to account for any differences in cognitive ability between the two diagnosis groups (TD and NDD).

### 2.3 Neuromelanin-Sensitive Magnetic Resonance Imaging

#### 2.3.1 MRI Acquisition

All participants were scanned on a 3T Prisma Fit scanner (Siemens, Erlangen, Germany), with a 32-channel head coil at Western University’s Centre for Functional and Metabolic Mapping. NM-MRI signal concentration was measured using a modified three-dimensional (3D) gradient-recalled echo with magnetization-transfer pulse (GRE-MT) NM-MRI image with the following parameters: repetition time (TR)=54ms; echo time (TE)=7.34ms; flip angle=16°; FoV=192×192; matrix=320×320; slice thickness=3mm; acquisition time=5:12min. A T1-weighted (T1w) image was acquired for NM-MRI image processing using a 3D magnetization prepared rapid acquisition gradient echo (MPRAGE) sequence with the following parameters: spatial resolution=1×1×1mm^3^; TR=2,300ms; TE=2.88ms; inversion time (TI)=900ms; flip angle=9°; acquisition time=5:00min; FoV=240×256; matrix=240×256; slice thickness=1mm; GRAPPA=2. Following acquisition, the NM-MRI images were visually inspected for motion artifacts affecting the midbrain.

#### 2.3.2 NM-MRI Preprocessing and Segmentation

Statistical Parametric Mapping (SPM12)^21^ in MATLAB (vR2022b) was used to preprocess NM-MRI scans. This allowed for voxelwise analyses in standardized MNI (Montreal Neurological Institute) space. NM-MRI images were first realigned to correct for motion using ‘SPM-Realign: Estimate and Reslice.’ Then, the realigned and estimated NM-MRI images were averaged using ‘SPM-ImCalc.’ The averaged NM-MRI image was then co-registered to the T1w image using ‘*SPM-Coregister: Estimate and Reslice’*. The realigned T1w image was segmented into different tissue types and spatially normalized to a standard MNI template using DARTEL.^22^ Using a 1mm full-width-at-half-maximum 3D Gaussian kernel with ‘SPM-Smooth’, the spatially normalized NM-MRI image was spatially smoothed. Lastly, the smoothed NM-MRI image was visually inspected relative to the template using ‘SPM-Check Reg.’

ITK-Snap (v. 3.8.0, www.itksnap.org) was leveraged to manually segment the SN in native MRI space. The crus cerebri (CC) and pontine tegmentum (PT) were also manually segmented as reference regions for contrast-to-noise ratio (CNR) analyses, as white-matter tracts are known to have marginal neuromelanin content^19^. The manual segmentation protocol followed previously described methods^19,23,24^ with reference to an anatomical MRI atlas of the mesencephalon^25^.

#### 2.3.3 Contrast-to-Noise Ratio (CNR) Calculations

To quantify the contrast between the signal of interest and background noise, manual tracking-based segmentations methods were leveraged.^26^ A contrast-to-noise (CNR) value corresponding to a single ROI is the most widely used metric in NM-MRI research; its formula has been adapted from prior work: [(signal – reference)/reference*100)], yielding a metric of neuromelanin percent signal change relative to reference regions^19^. Here, we use the terms NM-MRI % signal change and NM-MRI signal intensity interchangeably. SN masks were averaged bilaterally to account for intraindividual differences.

### 2.4 Statistical Analysis

Statistical analyses were performed using RStudio. Wilcoxon rank-sum tests and Fisher’s exact tests were conducted to examine demographic differences between diagnosis groups (TD, NDDs). Generalized linear models were run to examine NM-MRI signal intensity related to head-banging (SIB-head) and apathy.

A principal component analysis (PCA) was conducted to reduce dimensionality and identify underlying structure among the apathy-related BASC-3 extracted variables. The number of retained components was determined using eigenvalues >1, inspection of the scree plot, and interpretability of the component structure. Component loadings were examined following varimax rotation. Individual component scores were extracted and standardized for use in subsequent analyses.

To address our first aim, apathy component score was the dependent variable and head-banging severity was the independent variable. For our second aim, apathy score was the dependent variable and SN-NM-MRI signal intensity was the independent variable. Lastly, for our third aim, apathy score was the dependent variable and the interaction between head banging and SN-NM-MRI signal intensity was the independent variable. Sex, age, with diagnostic group as a factor, were adjusted for throughout all three aims. Continuous variables, including apathy scores, NM-MRI signal values, head-banging severity, and age, were standardized (mean = 0, SD = 1) prior to statistical analyses to improve comparability across measures and aid interpretation of model estimates.

## 3. RESULTS

### 3.1 Participant Characteristics

A total of 47 participants, 14 typically developing and 33 with neurodevelopmental disorders (ADHD, ASD), were included in this study (table 1). Approximately 64% of participants were male, and the average age of participants was 11 years old. All participants had complete BASC-3 data, while 70% had available NM-MRI data, and 68% had SIB data available. The neurodevelopmental disorders group demonstrated significantly higher BASC-3 sum scores on the items extracted to represent apathy compared to the typically developing group (NDD=2.72 ± 0.35, TD=2.38 ± 0.32, *p*<0.05).

**Table 1.** Participant demographics. Abbreviations: TD, typically developing; NDD, neurodevelopmental disorders (ADHD, ASD); (*) indicate statistically significant differences.

|  | TD (n=14) | NDD (n=33) | Total (n=47) | Contrasts |
| --- | --- | --- | --- | --- |
| Age, years (mean ± SD) | 9.75 ± 2.76 | 11.48 ± 2.70 | 10.96 ± 2.99 | W=152,<br>*p*=0.07 |
| Sex, (%) male | 7 (50) | 23 (70) | 30 (64) | OR=2.26,<br>*p*=0.32 |
| *Self-Injurious Behaviors (SIB)* |  |  |  |  |
| SIB-head=0, n (%) | 8 (57) | 18 (55) | 32 (55) |  |
| SIB-head severity (mean ± SD) | 0.20 ± 0.53 | 0.16 ± 0.37 | 0.17 ± 0.42 | W=112,<br>*p*=0.93 |
| SIB-head available, n (%) | 10 (71) | 22 (67) | 32 (68) |  |
| *Neuromelanin (NM)-MRI* |  |  |  |  |
| SN-NM-MRI CNR (% Signal change) | 12.65 ± 3.93 | 15.36 ± 3.58 | 14.38 ± 3.88 | W=75,<br>*p*=0.06 |
| SN-NM-MRI available, n (%) | 12 (86) | 21 (64) | 33 (70) |  |
| *The Behavior Assessment System for Children (BASC-3)* |  |  |  |  |
| BASC-3-Apathy (mean ± SD) | 2.38 ± 0.32 | 2.72 ± 0.35 | 2.62 ± 0.37 | W=109,<br>*p*<0.05* |
| *Full Scale IQ (FSIQ)* |  |  |  |  |
| FSIQ n (mean ± SD) | 103.2 ± 18.29 | 92.12 ± 13.61 | 95.19 ± 15.61 | W=175.5,<br>*p*=0.11 |
| FSIQ available, n (%) | 10 (71) | 26 (79) | 36 (77) |  |

### 3.2 Principal Component Analysis

Thirty-five items from the BASC-3 (table S1) were included in the PCA. A one-component model was determined to be favourable. This component (PC1) explained 47.3% of the total variance. Following varimax rotation, 16 item variables with factor loadings >0.50 were considered significant and retained in PC1 (table 2). PC1 scores were extracted and used to index apathy in subsequent analyses.

**Table 2.** PC1 item factor loadings >0.50.

| Item Number | Item | Factor Loading |
| --- | --- | --- |
| 2 | Plans well | 0.727 |
| 3 | Is a "self-starter" | 0.740 |
| 4 | Is usually chosen as a leader | 0.778 |
| 7 | Sets realistic goals | 0.767 |
| 9 | Shows interest in others' ideas | 0.575 |
| 15 | Stares blankly | 0.582 |
| 17 | Finds ways to solve problems | 0.752 |
| 21 | Has trouble making new friends | 0.561 |
| 22 | Gives good suggestions for solving problems | 0.629 |
| 26 | Shows basic emotions clearly | 0.546 |
| 28 | Organizes chores or other tasks well | 0.730 |
| 29 | Seems unaware of others | 0.624 |
| 30 | Prefers to be a leader | 0.518 |
| 32 | Makes friends easily | 0.600 |
| 34 | Congratulates others when good things happen to them | 0.626 |
| 35 | Is able to describe feelings accurately | 0.584 |

### 3.3 Association between apathy and heading-banging severity

Exploratory linear regression analyses were conducted to examine the relationship between apathy and head-banging severity, with sex, age, and diagnostic group as covariates. There were no significant findings of greater head-banging severity being associated with higher apathy scores (F(4,26)=0.72, *p*=0.59).

### 3.4 Association between apathy and SN-NM-MRI signal intensity

Next, we investigated the relationship between apathy and neuromelanin-sensitive MRI signal intensity within the substantia nigra. Overall, greater apathy was not found to be associated with altered NM-MRI signal (F(4,28)=1.38, *p*=0.27).

### 3.5 Heading-banging severity as a moderator of the relationship between apathy and SN-NM-MRI intensity

Lastly, to test whether repetitive self-injurious behaviour to the head influenced the relationship between apathy and NM-MRI signal intensity, an exploratory moderation analysis was conducted including the interaction between NM-MRI signal intensity in the substantia nigra and head-banging severity. A significant interaction effect was observed (β=1.27, *p*=0.01; F(5,18)=2.78, *p*=0.044), indicating that the association between apathy and NM-MRI signal intensity varied as a function of head-banging severity. Specifically, higher levels of head-banging severity were associated with a stronger positive relationship between apathy and SN-NM-MRI signal (fig 1A). These findings suggest that repetitive head impacts may modify the relationship between motivational symptoms and dopaminergic system integrity. Additionally, diagnostic group was a significant predictor of apathy (β=0.75, *p*=0.04), where a Fisher’s exact test revealed that apathy was greater in the NDD group compared to the TD group (NDD: 0.27, TD: −0.60, W=108, *p*<0.05; fig 1B).

**Figure 1.**
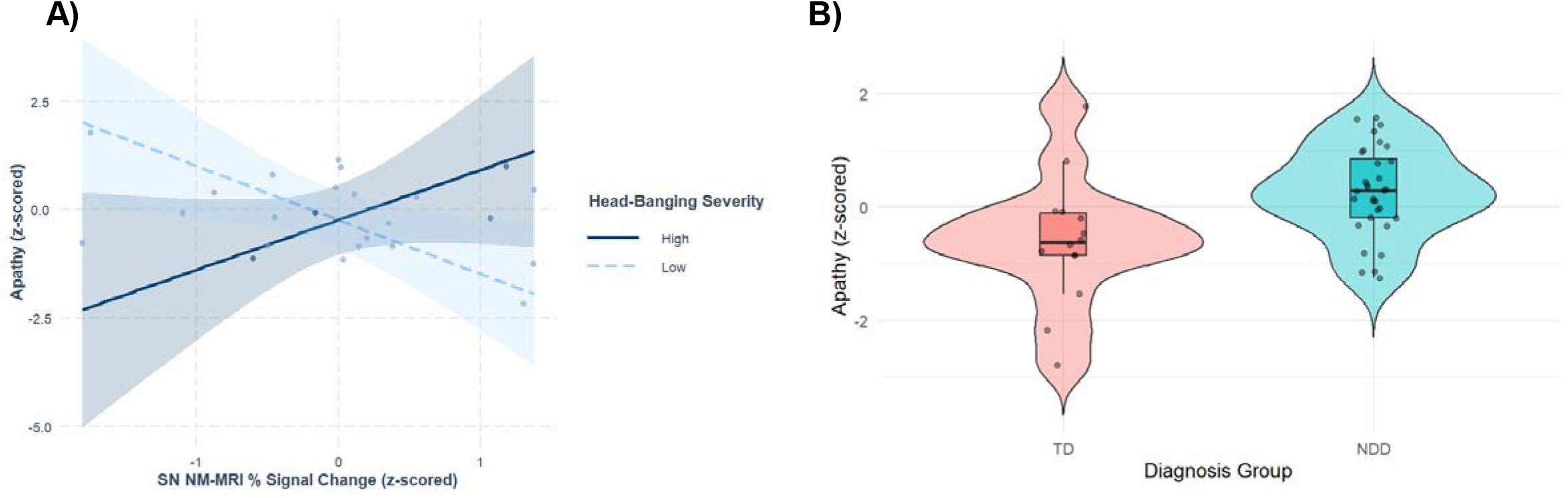
A) Interaction between SN NM-MRI % signal change and head-banging severity in predicting apathy. Regression lines illustrate the association between standardized SN NM-MRI % signal change and standardized apathy PC1 scores at low (−1 SD; dashed line) and high (+1 SD; solid line) levels of head-banging severity. Shaded regions represent 95% confidence intervals. Higher SN NM-MRI % signal change was associated with greater apathy severity at high levels of head-banging severity, whereas the association was reduced or reversed at low levels of head-banging severity. **B) Main effect of diagnosis group on apathy**. Violin plots depict the distribution density of apathy PC1 z-scores for typically developing (TD) participants and participants with neurodevelopmental disorders (NDD). Boxplots represent the median and interquartile range, while individual data points indicate participant-level observations. Higher values reflect greater apathy severity.

## 4. DISCUSSION

The present exploratory study investigated associations between apathy, repetitive head-banging behaviour, and substantia nigra neuromelanin-sensitive MRI signal in children and adolescents with neurodevelopmental disorders. Using harmonized BASC-3 items, we identified a principal component (PC1) reflecting motivational, social-emotional, and goal-directed behavioural characteristics consistent with apathy-related symptomatology. While direct associations between apathy and head-banging severity or NM-MRI signal intensity were not observed independently, a significant interaction emerged between head-banging severity and substantia nigra NM-MRI signal intensity in predicting apathy. Together, these findings provide preliminary evidence that repetitive behaviour causing self-injury to the head may influence the relationship between motivational symptoms and dopaminergic system integrity during development.

Consistent with prior literature, youth with NDDs demonstrated significantly higher apathy-related scores compared to typically developing peers.^27^ Many of the retained BASC-3 items reflected behavioural activation, planning, emotional awareness, and social reciprocity, domains frequently implicated in apathy.^28^ Given the absence of a validated pediatric apathy instrument, the current findings suggest that dimensional approaches using existing behavioural measures may provide a feasible strategy for indexing apathy-related constructs across development. Importantly, the PCA-derived component demonstrated a coherent one-factor structure, supporting the conceptual overlap among these motivational and socioemotional behaviours.

Contrary to our initial hypothesis, head-banging severity alone was not significantly associated with apathy scores. Similarly, substantia nigra NM-MRI % signal change was not independently associated with apathy. However, these null findings should be interpreted cautiously given the modest sample size and limited statistical power. Apathy is multifaceted and influenced by a combination of developmental, behavioural, and neurobiological factors that may not be detectable through linear associations in small pediatric samples. Furthermore, the restricted range of head-banging severity observed in this cohort may have limited sensitivity to detect direct effects.

Although the biological interpretation of increased NM-MRI signal intensity in this context of head-banging and apathy remains unclear, alterations in neuromelanin-sensitive signal in the substantia nigra may reflect developmental or compensatory changes within catecholaminergic systems following repetitive head impacts. These findings align with broader evidence implicating dopaminergic dysfunction in motivational impairment following traumatic brain injury. Experimental and clinical studies have shown that TBI can disrupt dopaminergic transmission within nigrostriatal and mesocorticolimbic pathways involved in reward processing, behavioural activation, and goal-directed behaviour, and such alterations may contribute to persistent neuropsychiatric symptoms, including apathy.^29,30^ Similar patterns of dopaminergic dysfunction have also been reported in adults exposed to repetitive head impacts, including former American football players.^31^ The present findings provide preliminary evidence that repetitive head-banging behaviour in youth with neurodevelopmental disorders may be associated with early alterations in catecholaminergic systems linked to motivational functioning and potentially long-term neuropsychiatric risk.

The use of NM-MRI in pediatric populations represents an emerging area of neuroimaging research. NM-MRI provides a non-invasive *in vivo* proxy of catecholaminergic neuron integrity within brainstem structures such as the substantia nigra, which plays a central role in reward processing, behavioural activation, and goal-directed behaviour. While NM-MRI has been increasingly studied in neurodegenerative disorders, relatively little work has examined its utility in developmental populations or in relation to repetitive self-injurious behaviours.^32^ The present findings therefore provide preliminary support for the feasibility of combining behavioural apathy measures with NM-MRI to investigate motivational circuitry in youth with NDDs.

Several limitations should be considered. First, the study was exploratory and limited by a small sample size, particularly for analyses involving NM-MRI and self-injurious behaviour data, reducing statistical power and limiting generalizability. Second, the apathy construct was derived from harmonized BASC-3 items rather than a validated pediatric apathy instrument. Although the PCA supported a coherent factor structure, further psychometric validation is needed. Third, head-banging severity was based on available behavioural measures and may not fully capture cumulative repetitive head impact exposure. Finally, the cross-sectional design limits conclusions regarding causality or developmental trajectories.

Despite these limitations, this study provides initial evidence supporting a potential interaction between repetitive head-banging behaviour and dopaminergic system integrity in relation to apathy-related symptoms in youth with NDDs. Future studies with larger longitudinal cohorts are needed to validate pediatric measures of apathy, better characterize repetitive head injury exposure, and determine whether NM-MRI biomarkers may help identify children at risk for motivational and neuropsychiatric difficulties following repetitive head impacts.

## Data Availability

All data produced in the present study are available upon reasonable request to the authors

## Data availability

Data will be available from the corresponding author upon request.

## Acknowledgements

No acknowledgements.

## Author contributions

R.M. and E.G.D. conceptualised the study, S.A.A. acquired and processed the neuroimaging data with oversight from E.G.D., K.R. administered the BASC-3, R.M. wrote the first manuscript draft, E.G.D., S.A.A., and K.R. provided input on the analytic approach and revised the manuscript. All authors reviewed the final manuscript.

## REFERENCES

1. Husain M, Roiser JP. Neuroscience of apathy and anhedonia: a transdiagnostic approach. Nat Rev Neurosci. 2018;19(8):470–484. doi:10.1038/s41583-018-0029-9

2. Levy R, Dubois B. Apathy and the functional anatomy of the prefrontal cortex-basal ganglia circuits. Cereb Cortex. 2006;16(7):916–928. doi:10.1093/cercor/bhj043

3. Marin RS. Apathy: a neuropsychiatric syndrome. J Neuropsychiatry Clin Neurosci. 1991;3(3):243–254. doi:10.1176/jnp.3.3.243

4. Arnould, A., Rochat, L., Azouvi, P. et al. A Multidimensional Approach to Apathy after Traumatic Brain Injury. Neuropsychol Rev 23, 210–233 (2013). 10.1007/s11065-013-9236-3

5. Tierney, S. M., Woods, S. P., Weinborn, M., & Bucks, R. S. (2018). Real-world implications of apathy among older adults: Independent associations with activities of daily living and quality of life. Journal of Clinical and Experimental Neuropsychology, 40, 895–903. https://www.tandfonline.com/doi/abs/10.108013803395.2018.1444736

6. Tsai CF, Hwang WS, Lee JJ, et al. Predictors of caregiver burden in aged caregivers of demented older patients. BMC Geriatr. 2021;21(1):59. Published 2021 Jan 14. doi:10.1186/s12877-021-02007-1

7. Baugh, C. M., Stamm, J. M., Riley, D. O., Gavett, B. E., Shenton, M. E., Lin, A., et al. (2012). Chronic traumatic encephalopathy: neurodegeneration following repetitive concussive and subconcussive brain trauma. Brain Imaging and Behavior, 6(2), 244–254.

8. Maietta JE, Iverson GL and Cook NE (2025) Lifetime history of concussion among children and adolescents with attention-deficit/hyperactivity disorder: examining differences stratified by age, medication status, and parent-reported severity. Front. Neurol. 15:1487909. doi: 10.3389/fneur.2024.1487909

9. DiGuiseppi, C., Levy, S. E., Sabourin, K. R., Soke, G. N., Rosenberg, S., Lee, L. C., Moody, E., & Schieve, L. A. (2018). Injuries in Children with Autism Spectrum Disorder: Study to Explore Early Development (SEED). Journal of autism and developmental disorders, 48(2), 461–472. 10.1007/s10803-017-3337-4

10. Pakyurek, M., Badawy, M., Ugalde, I. T., Ishimine, P., Chaudhari, P. P., McCarten-Gibbs, K., Nobari, O., Kuppermann, N., & Holmes, J. F. (2022). Does attentiondeficit/hyperactivity disorder increase the risk of minor blunt head trauma in children?. Journal of child and adolescent psychiatric nursing : official publication of the Association of Child and Adolescent Psychiatric Nurses, Inc, 35(4), 356–361. 10.1111/jcap.12390

11. Dimian, A.F., Botteron, K.N., Dager, S.R. et al. Potential Risk Factors for the Development of Self-Injurious Behavior among Infants at Risk for Autism Spectrum Disorder. J Autism Dev Disord 47, 1403–1415 (2017). 10.1007/s10803-017-3057-9

12. Summers, J., Shahrami, A., Cali, S., D’Mello, C., Kako, M., Palikucin-Reljin, A., Savage, M., Shaw, O., & Lunsky, Y. (2017). Self-Injury in Autism Spectrum Disorder and Intellectual Disability: Exploring the Role of Reactivity to Pain and Sensory Input. Brain sciences, 7(11), 140. 10.3390/brainsci7110140

13. Meiling, J. B., Schulze, D. R., Hines, E., Hassett, L. C., & Esterov, D. (2022). Traumatic Brain Injury After Music-Associated Head Banging: A Scoping Review. Archives of rehabilitation research and clinical translation, 4(3), 100192. 10.1016/j.arrct.2022.100192

14. Hof, P. R., Knabe, R., Bovier, P., & Bouras, C. (1991). Neuropathological observations in a case of autism presenting with self-injury behavior. Acta neuropathologica, 82(4), 321–326. 10.1007/BF00308819

15. Hewitt, S. R. C., Habicht, J., Bowler, A., Lockwood, P. L., & Hauser, T. U. (2024). Probing apathy in children and adolescents with the Apathy Motivation Index-Child version. Behavior research methods, 56(4), 3982–3994. 10.3758/s13428-023-02184-4

16. Bourke, N. J., Demarchi, C., De Simoni, S., Samra, R., Patel, M. C., Kuczynski, A., Mok, Q., Wimalasundera, N., Vargha-Khadem, F., & Sharp, D. J. (2022). Brain volume abnormalities and clinical outcomes following paediatric traumatic brain injury. Brain : a journal of neurology, 145(8), 2920–2934. 10.1093/brain/awac130

17. Stephens, J. A., Salorio, C. F., Gomes, J. P., Nebel, M. B., Mostofsky, S. H., & Suskauer, S. J. (2017). Response Inhibition Deficits and Altered Motor Network Connectivity in the Chronic Phase of Pediatric Traumatic Brain Injury. Journal of neurotrauma, 34(22), 3117–3123. 10.1089/neu.2017.5081

18. Sood, N. T., Godfrey, C., Chavez Arana, C., Anderson, V., & Catroppa, C. (2023). Paediatric traumatic brain injury and the dysregulation profile: The mediating role of decision-making. Neuropsychological rehabilitation, 33(3), 440–453. 10.1080/09602011.2022.2025861

19. Cassidy, C. M., Zucca, F. A., Girgis, R. R., Baker, S. C., Weinstein, J. J., Sharp, M. E., Bellei, C., Valmadre, A., Vanegas, N., Kegeles, L. S., Brucato, G., Kang, U. J., Sulzer, D., Zecca, L., Abi-Dargham, A., & Horga, G. (2019). Neuromelanin-sensitive MRI as a noninvasive proxy measure of dopamine function in the human brain. Proceedings of the National Academy of Sciences of the United States of America, 116(11), 5108–5117. 10.1073/pnas.1807983116

20. Kaiser, S., & Schlagenhauf, F. (2021). Brain reward systems and apathy. In A. Aleman & K. L. Lanctôt (Eds.), Apathy: Clinical and neuroscientific perspectives from neurology and psychiatry (pp. 191–211). Oxford University Press. 10.1093/med/9780198841807.003.0011

21. Friston, K. J., et al. (1995). “Statistical Parametric Maps in Functional Imaging: A General Linear Approach.” Human Brain Mapping.

22. Ashburner, J. (2007). A fast diffeomorphic image registration algorithm. NeuroImage, 38(1), 95–113.

23. Eapen M, Zald DH, Gatenby JC, Ding Z, Gore JC. Using high-resolution MR imaging at 7T to evaluate the anatomy of the midbrain dopaminergic system. AJNR Am J Neuroradiol. 2011;32(4):688–694. doi:10.3174/ajnr.A2355

24. Betts MJ, Cardenas-Blanco A, Kanowski M, Jessen F, Düzel E. In vivo MRI assessment of the human locus coeruleus along its rostrocaudal extent in young and older adults. Neuroimage. 2017;163:150–159. doi:10.1016/j.neuroimage.2017.09.042

25. Duvernoy HM. The human brain stem and cerebellum: surface, structure, vascularization, and three-dimensional sectional anatomy, with MRI. Springer Science & Business Media; 2012.

26. Lv Q, Wang X, Lin P, Wang X. Neuromelanin-sensitive magnetic resonance imaging in the study of mental disorder: A systematic review. Psychiatry Res Neuroimaging. 2024;339:111785. doi:10.1016/j.pscychresns.2024.111785

27. Torrente F, Lischinsky A, Torralva T, López P, Roca M, Manes F. Not always hyperactive? Elevated apathy scores in adolescents and adults with ADHD. J Atten Disord. 2011;15(7):545–556. doi:10.1177/1087054709359887

28. Ang, Y. S., Lockwood, P., Apps, M. A., Muhammed, K., & Husain, M. (2017). Distinct Subtypes of Apathy Revealed by the Apathy Motivation Index. PloS one, 12(1), e0169938. 10.1371/journal.pone.0169938

29. Chen, Y. H., Huang, E. Y., Kuo, T. T., Miller, J., Chiang, Y. H., & Hoffer, B. J. (2017). Impact of Traumatic Brain Injury on Dopaminergic Transmission. Cell transplantation, 26(7), 1156–1168. 10.1177/0963689717714105

30. Chong TT, Husain M. The role of dopamine in the pathophysiology and treatment of apathy. Prog Brain Res. 2016;229:389–426. doi:10.1016/bs.pbr.2016.05.007

31. van Amerongen S, Peskind ER, Tripodis Y, et al. Catecholamine Dysregulation in Former American Football Players: Findings From the DIAGNOSE CTE Research Project. Neurology. 2025;104(10):e213584. doi:10.1212/WNL.0000000000213584

32. Nuraini N, Appling C, Ferguson BJ, et al. A Preliminary Investigation of Dopamine Transporter Binding Abnormalities in Individuals With Autism Spectrum Disorder. Autism Res. 2026;19(1):e70144. doi:10.1002/aur.70144

