## Supplemental Table 1 for "Exploring the Relationship Between Apathy, Dopaminergic Signal, and Head Injury in Neurodevelopmental Disorders"

| Study Item # | Child ITEM # | Child Form | Adol ITEM # | Adolescent Form |
| --- | --- | --- | --- | --- |
| 1 | 9 | Worries | 4 | Worries |
| 2 | 16 | Plans well | 15 | Plans well |
| 3 | 18 | Is a "self-starter | 17 | Is a "self-starter" |
| 4 | 29 | Is usually chosen as a leader | 29 | Is usually chosen as a leader |
| 5 | 34 | Cries easily | 89 | Cries easily |
| 6 | 36 | Avoids exercise or other physical activity | 24 | Avoids exercise or other physical activity |
| 7 | 37 | Sets realistic goals | 36 | Sets realistic goals |
| 8 | 38 | Worries about things that cannot be changed | 135 | Worries about things that cannot be changed |
| 9 | 53 | Shows interest in others' ideas | 51 | Shows interest in others' ideas |
| 10 | 66 | Needs to be reminded to brush teeth | 60 | Needs to be reminded to brush teeth |
| 11 | 71 | Takes a step-by-step approach to work | 64 | Takes a step-by-step approach to work |
| 12 | 60 | Is sad | 19 | Is sad |
| 13 | 86 | Accepts things as they are | 82 | Accepts things as they are |
| 14 | 87 | Quickly joins group activities | 83 | Quickly joins group activities |
| 15 | 88 | Stares blankly | 84 | Stares blankly |
| 16 | 90 | Cleans up after self | 86 | Cleans up after self |
| 17 | 95 | Finds ways to solve problems | 173 | Finds ways to solve problems |
| 18 | 102 | Likes to talk about his or her day | 8 | Likes to talk about his or her day |
| 19 | 107 | Worries about what teachers think | 99 | Worries about what teachers think |
| 20 | 109 | Starts conversations | 101 | Starts conversations |
| 21 | 111 | Has trouble making new friends | 113 | Has trouble making new friends |
| 22 | 120 | Gives good suggestions for solving problems | 132 | Gives good suggestions for solving problems |
| 23 | 126 | Isolates self from others | 33 | Isolates self from others |
| 24 | 128 | Worries about making mistakes | 120 | Worries about making mistakes |
| 25 | 138 | Is overly emotional | 44 | Is overly emotional |
| 26 | 139 | Shows basic emotions clearly | 159 | Shows basic emotions clearly |
| 27 | 142 | Makes decisions easily | 136 | Makes decisions easily |
| 28 | 149 | Organizes chores or other tasks well | 143 | Organizes chores or other tasks well |
| 29 | 152 | Seems unaware of others | 150 | Seems unaware of others |
| 30 | 155 | Prefers to be a leader | 171 | Prefers to be a leader |
| 31 | 160 | Says, "I'm afraid I will make a mistake | 153 | Says, "I'm afraid I will make a mistake" |
| 32 | 163 | Makes friends easily | 165 | Makes friends easily |
| 33 | 173 | Is highly motivated to succeed | 148 | Is highly motivated to succeed |
| 34 | 174 | Congratulates others when good things happen to them | 170 | Congratulates others when good things happen to them |
| 35 | 165 | Is able to describe feelings accurately | 142 | Is able to describe feelings accurately |

Table S1. BASC-3-Apathy. Items from the BASC-3 Child and Adolescent forms extracted to reflect apathy and leveraged in a principal component analysis (PCA).
